# Association of age at menarche and menopause, reproductive lifespan, and stroke among Chinese women: Results from a national cohort study

**DOI:** 10.1101/2023.05.23.23290429

**Authors:** Lu Chen, Zhen Hu, Xin Wang, Congyi Zheng, Xue Cao, Jiayin Cai, Yixin Tian, Runqing Gu, Linfeng Zhang, Zengwu Wang

## Abstract

**Background:** The association between age of menarche, age of menopause, reproductive lifespan and risk of stroke in Chinese women remains unclear and requires further clarification.

**Methods:** A stratified multi-stage random sampling method was used to select participants at baseline in 2012-2015. The participants’ basic information was collected through a standardized questionnaire by professional investigator and examined by trained medical personnel. Follow-up was completed in 2019 to collect the stroke events. The Cox proportional hazards models were used to evaluate hazard ratios between reproductive factors and stroke risk.

**Results:** Overall, 11,256 women (5,155 non-menopausal women and 6,101 menopausal women) were included (mean [standard deviation] age, 55.2 [12.9] years). The risk was highest in women with menarche at age ≥17 years (HR, 1.290; 95%CI, 0.959∼1.733) and with reproductive lifespan ≤28 years (HR, 1.643; 95%CI, 1.041∼2.595). Age at menarche was positively associated with risk of stroke (HR, 1.086; 95%CI, 1.006∼1.172). There was a negative association between age at menopause and stroke risk in women with 2 live births (HR, 0.897; 95%CI, 0.834∼0.964). Reproductive lifespan was negatively associated with risk of stroke (HR, 0.963; 95%CI, 0.946∼1.027). Subgroup analysis also showed that association between age at menarche, reproductive lifespan and stroke events.

**Conclusions:** Chinese women with late age at menarche and shorter reproductive lifespan have higher risk of stroke according to a large prospective study.

## 1. Intruduction

Stroke is the third most common cause of death in women and men, after heart disease and cancer, along with being one of the principal causes of disability [1]. Studies demonstrate that there are some important differences between the sexes that younger females are better protected from stroke than men [2]. This difference reverses as age advances in females and menopause ensues, resulting in higher incidence of stroke in females [2]. In addition, women report nonconventional symptoms of stroke more frequently than men which leads to delays in seeking medical attention, diagnosis, and accessing acute stroke services. Therefore, increasing awareness of the specific risk factors and warning signs of stroke that are unique to women may help reduce the total burden of cerebrovascular disease. Menopause increases the stroke risk in females as corroborative fromepidemiological study and animal studies [3, 4]. This is thought to be because of the protective effects of ovarian hormones [5]. Hormonal factors, such as the decline in oestrogen and the relative increase in androgens in postmenopausal women, are believed to increase the risk and incidence of ischaemic stroke in this population [6]. This suggests that the increased risk of stroke in women may be related to reproductive factors, including age at menarche, age at menopause, and reproductive lifespan. Menarche is the beginning, and menopause is the end of a woman’s reproductive timeline, and the reproductive years, which is the interval between menarche and menopause, is a woman’s natural reproductive window. Due to exposure to different hormonal levels, early or late onset of these events may be associated with increased risk of many chronic health problems, in particular cardiovascular and cerebrovascular disease, such as stroke. Clarifying this effect could provide important information on new plausible paradigms to prevent stroke and ischemic vascular disease. However, the association between duration of reproductive factors and stroke risk has not been investigated thoroughly, which needs to be validated in large, long-term cohort studies.

Based on a national cohort study in China, we investigate the associations between age at menarche and menopause, reproductive lifespan, and the stroke risk and examine the relationship in subgroups amongst Chinese women.

## 2. Materials and Methods

### 2.1 Study design and participants

From October 2012 to December 2015, our team carried out a representative large-scale cross-sectional survey of cardiovascular diseases (CVD) in China. A stratified multi-stage random sampling method was used to select about 500,000 participants aged ≥15 years from 262 districts and counties in 31 provinces in Chinese mainland. Demographic characteristics, lifestyle risk factors, drug therapy and female reproductive characteristics were collected. In the secondary sampling, simple random sampling method was adopted to select 33 districts and counties in the eastern, central and western regions respectively and stratified according to urban and rural areas. Blood samples for investigation of fasting plasma glucose (FPG), total cholesterol (TC), triglycerides (TG), low-density lipoprotein cholesterol (LDL-C), and high-density lipoprotein cholesterol (HDL-C) levels were collected from randomly selected participants aged ≥ 35 years from eligible areas. 30,036 subjects who completed the baseline survey were followed up in 2018-2019. 5,323 patients were lost to follow-up within 5 years, and the total follow-up rate was 82.3%. After excluding 619 subjects with a history of stroke, 10,665 male participants, and 2,173 postmenopausal women with incomplete information or errors, a total of 11,256 women were included in the final analysis (Figure 1). Obtain written informed consent from each participant. The Ethics Committee of Fuwai Hospital (Beijing, China) approved the study. This study follows STROBE guidelines (Strengthening the Reporting of Observational Studies in Epidemiology; Supplemental Material) [7].

**Figure 1.**
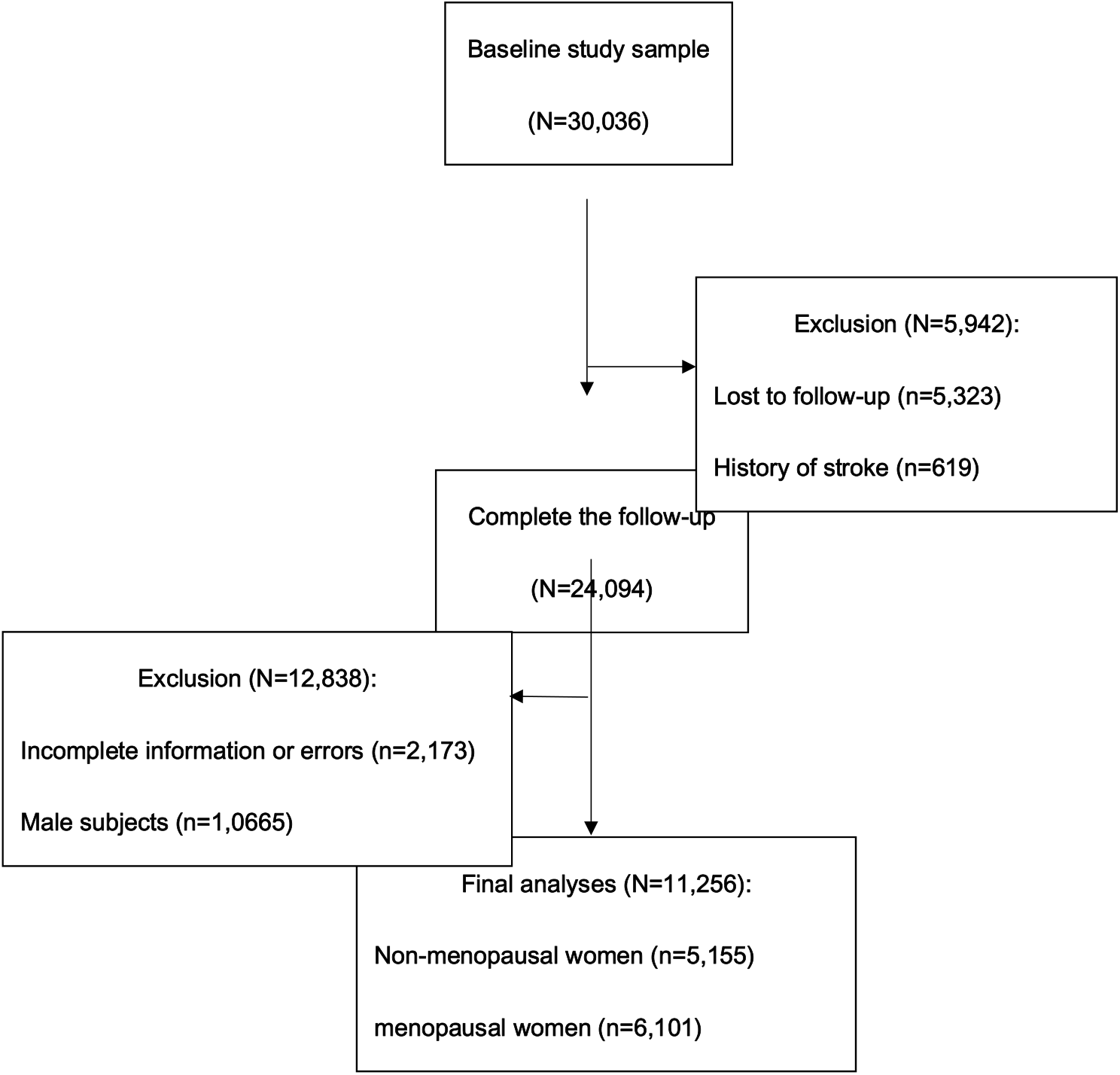
Flowchart of inclusion and exclusion of study participants. A schematic overview of the subject inclusion and exclusion.

### 2.2 Baseline Measurement and data collection

Information on demographic characteristics, lifestyle risk factors, medical history and female reproductive characteristics was collected by professional investigator using standardized questionnaires. Height was measured using a standard right-angle meter and weight was measured using the Omron Body Fat Scale (V-body HBF-371, Omron, Japan) with participants wearing light clothing and barefoot. Blood pressure (BP) was measured in the right upper arm after the subject rested for at least 5 minutes in the sitting position using an Omron HBP-1300 professional portable sphygmomanometer (Omron, Kyoto, Japan). Laboratory tests were performed on venous blood samples taken after at least 8 hours of overnight fasting. The levels of FPG, TC, TG, LDL-C and HDL-C were determined by the Central Core Laboratory (Beijing Adicam Clinical Laboratory Co., LTD., Beijing, China).

### 2.3 Follow-up and outcome measures

During the follow-up phase, the occurrence of stroke events was tracked through interviewing participants or their agents, or through telephone or mail questionnaires, and medical records were further checked for reconfirmation. The participants’ stroke events were initially recorded by local investigators, then the hospital records were reviewed by the Central Evaluation Committee of Fuwai Hospital (Beijing, China) to determine the final diagnosis. Stroke (ICD10 code I60–I61 and I63–I64) events are defined as an acute cerebrovascular disease caused by stenosis, occlusion or rupture of an artery in the brain due to various predisposing factors, resulting in an acute cerebral blood circulation disorder and limited or diffuse cerebral deficits, was divided into two types: ischaemic stroke and haemorrhagic stroke.

### 2.4 Variable definition

At baseline, each participant was asked about their age at the onset of their first period and menopause, which were recorded as age at menarche and age at menopause. According to the World Health Organization, menopausal status was defined as the cessation of menstruation for at least 12 months. Reproductive lifespan was measured by subtracting the age at menarche from the age at menopause.

Body mass index (BMI) was calculated by dividing weight (kg) by height squared (m^2^). BMI between 24.0∼27.9 kg/m^2^ was defined as overweight, and BMI≥28.0 kg/m^2^ as obese [8]. Alcohol drinking was defined as consuming alcoholic beverages at least once a week in the past month. Smoking was defined as having smoked at least 20 packs of cigarettes in a lifetime and still smoking [9]. Diabetes was defined as FBG ≥7.0 mmol/dL or a doctor’s diagnosed of diabetes or taking hypoglycemic drugs within 2 weeks [10]. Dyslipidemia as TG≥ 2.26mmol /L or TC≥ 6.22mmol /L or HDL-C<1.04 mmol/L or LDL-C≥4.14 mmol/L or previous use of lipid-lowering drugs or previous diagnosis of dyslipidemia according to the Guidelines for Dyslipidemia in China [11].

Drug treatment was defined as taking any kind of anti-hypertensive, hypoglycemic, or anti-hyperlipidemia drugs. Comorbidity were any combination of two or more of three diseases: hypertension, diabetes and dyslipidemia [12]. Parity was defined as the number of live births a woman gives birth to, whether or not the baby dies after delivery [13].

### 2.5 Statistical analysis

Basic characteristics of participants were expressed as mean and standard deviation (SD) of the normally distributed data or as a proportion of the categorical data. The χ^2^ test and variance analysis were used to compare variables among groups. Logistic linear regression model was used to explore the trend of variables. Multivariable Cox proportional hazard model was used to evaluate hazard ratios (HRs) and 95% confidence intervals (CIs) for stroke events associated with age at menarche (classified as≤13, 14, 15, 16, and≥17 years) with 15 years as the reference group, age at menopause (classified as≤44, 45-46, 47-48, 49-50, and ≥51 years) with 47-48 years as the reference group, and reproductive lifespan (classified as≤28, 29-31, 32-34, 35-37, and ≥38 years) with 32-34 years as the reference group.

This study further investigated the associations between stroke and menarche by adjusting for age at recruitment (continuous), BMI (continuous), region (urban or rural), marital status (unmarried/widowed, married/cohabiting), education level (elementary or below, junior high school, high school or above), alcohol drinking (yes or no), smoking (yes or no), drug treatment (yes or no), comorbidity (yes or no), family history of stroke (yes or no), menstrual status(yes or no), age at menarche (continuous), reproductive lifespan (continuous), age at menopause (continuous), contraceptive use (yes or no), breastfeeding experience (yes or no) and parity (0-1, 2, ≥3). We also examined the stroke risk in subgroups of women defined by region, age at recruitment, marital status, BMI,education level, comorbidity, drug treatment, alcohol drinking status, and family history of stroke, use of contraceptives, parity. R 3.6.2 software were used for analyses. The threshold of statistical significance was *P*<0.05.

## 3. Results

**Table 1, 2, and 3** respectively presented the characteristics of age at menarche and menopause, and reproductive lifespan of study participants. The mean (SD) age of participants was 55.2 (12.9) years and menopausal women was 63.4 (9.9) years. The mean age at menarche, age at menopause, reproductive lifespan and BMI and parity were 15.3 (1.8) years, 48.6 (3.6) years, 32.9 (3.9) years, 24.6 (3.6) kg/m^2^ and 2.4 (2.3), respectively. Those who smoked, drank, used contraceptives and had breastfeeding experience accounted for 2.5%, 6.9%, 9.2% and 95.4%, respectively.

Age at menarche was positively associated with stroke events in Model 1, Model 2 and Model 3, while reproductive lifespan was negatively associated, age at menopause was not significantly. Model 3 showed that the HRs (95%CIs) were 0.898 (0.596∼1.353), 12 0.957 (0.647∼1.414), 1.000 (reference), 1.202 (0.854∼1.691), and 1.290 (0.959∼1.733) 13 for women with age at menarche ≤13, 14, 15 (reference), 16, and ≥17 years, respectively; 14 1.551 (0.893∼2.690), 1.280 (0.720∼2.250), 1.000 (reference), 1.016 (0.632∼1.633), and 15 1.190 (0.699∼1.883) for women with age at menopause ≤44, 45-46, 47-48 (reference), 16 49-50, and ≥51 years; and 1.643 (1.041∼2.595), 1.225 (0.780∼1.920), 1.000 (reference), 17 1.099 (0.694∼1.711), and 0.980 (0.619∼1.796) for women with reproductive lifespan ≤28, 29-31, 32-34 (reference), 35-37, and ≥38. The risk was highest in women with menarche at age ≥17 years and with reproductive lifespan ≤28 years **(Figure 2**).

**Figure 2.**
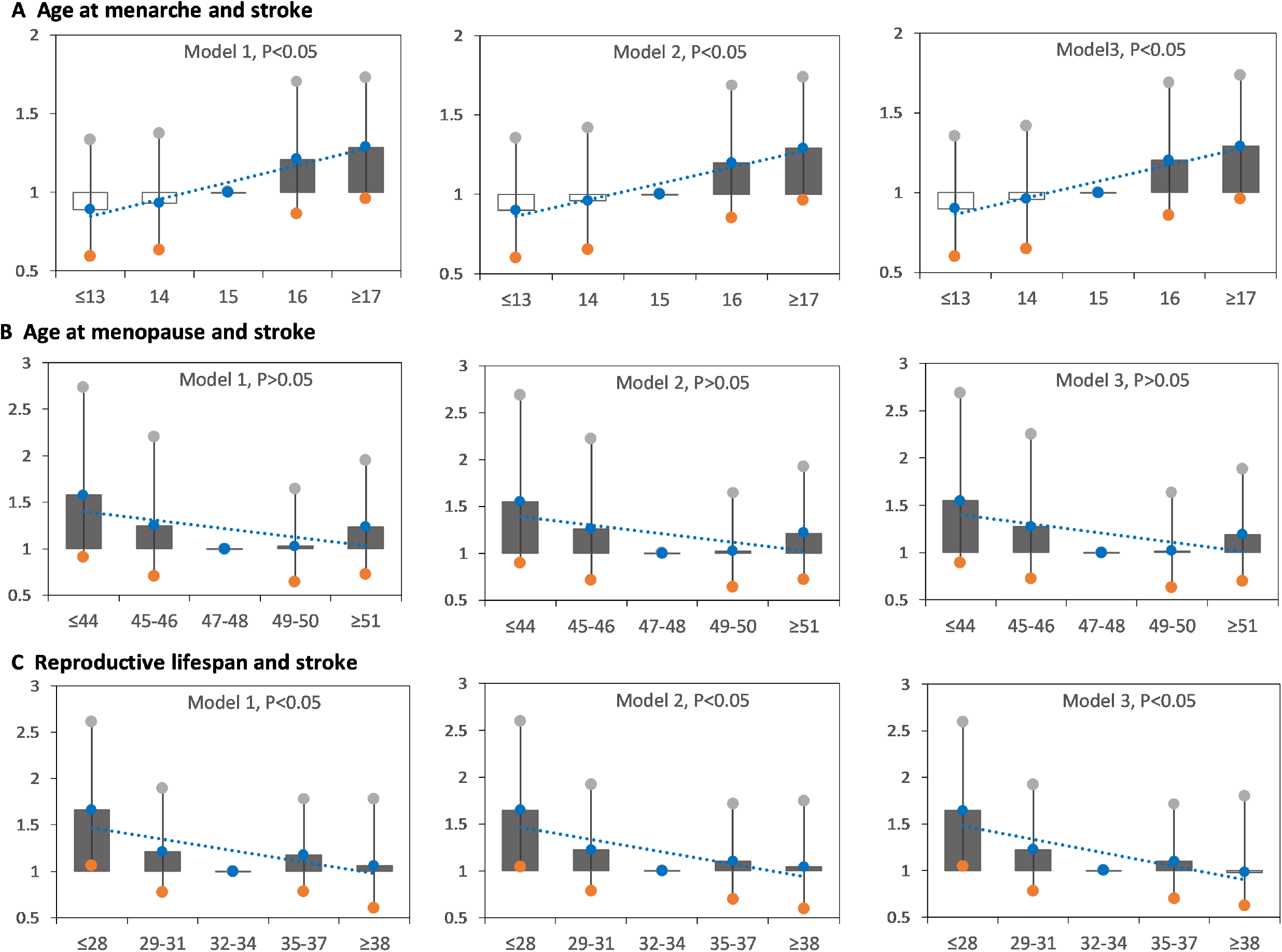
Hazard ratios (HRs) for stroke by age at menarche, age at menopause, and reproductive lifespan among women. Model A1, model B1, and model C1: adjusted for age at recruitment. Model A2, model B2, and model C2: model A1, model B1, and model C1 plus region, marital status, body mass index, education level, alcohol drinking, smoking, comorbidity, pharmacological treatment, family history of stroke.Model A3: model A2 plus menstrual status, contraceptive use status, and breastfeeding experience, parity; Model B3: model B2 plus age at menarche, reproductive lifespan, contraceptive use status, and breastfeeding experience, parity; Model C3: model C2 plus age at menarche, age at menopause, contraceptive use status, and breastfeedingexperience, parity.

After adjustment, the HR (95%CI) of stroke risk for age at menarche, age at menopause and reproductive lifespan was 1.086 (1.006∼1.172), 0.986 (0.946∼1.027) and 0.963 (0.929∼0.998) per year. Subgroup analyses showed that age at menarche was positively associated with risk of stroke among women aged 60-70 years old, with low BMI, low education level, no alcohol consumption, and no previous contraceptive use. Reproductive lifespan was negatively associated with risk of stroke among women who were married, 60-70 years of age, low BMI, comorbidities, pharmacological treatment, not consume alcohol, not previous contraceptive use, and 2 live births. There also was a negative association between age at menopause and stroke risk in women with 2 live births, with an adjusted HR (95%CI) of 0.897 (0.834∼0.964) per year **(Figure 3)**.

**Figure 3.**
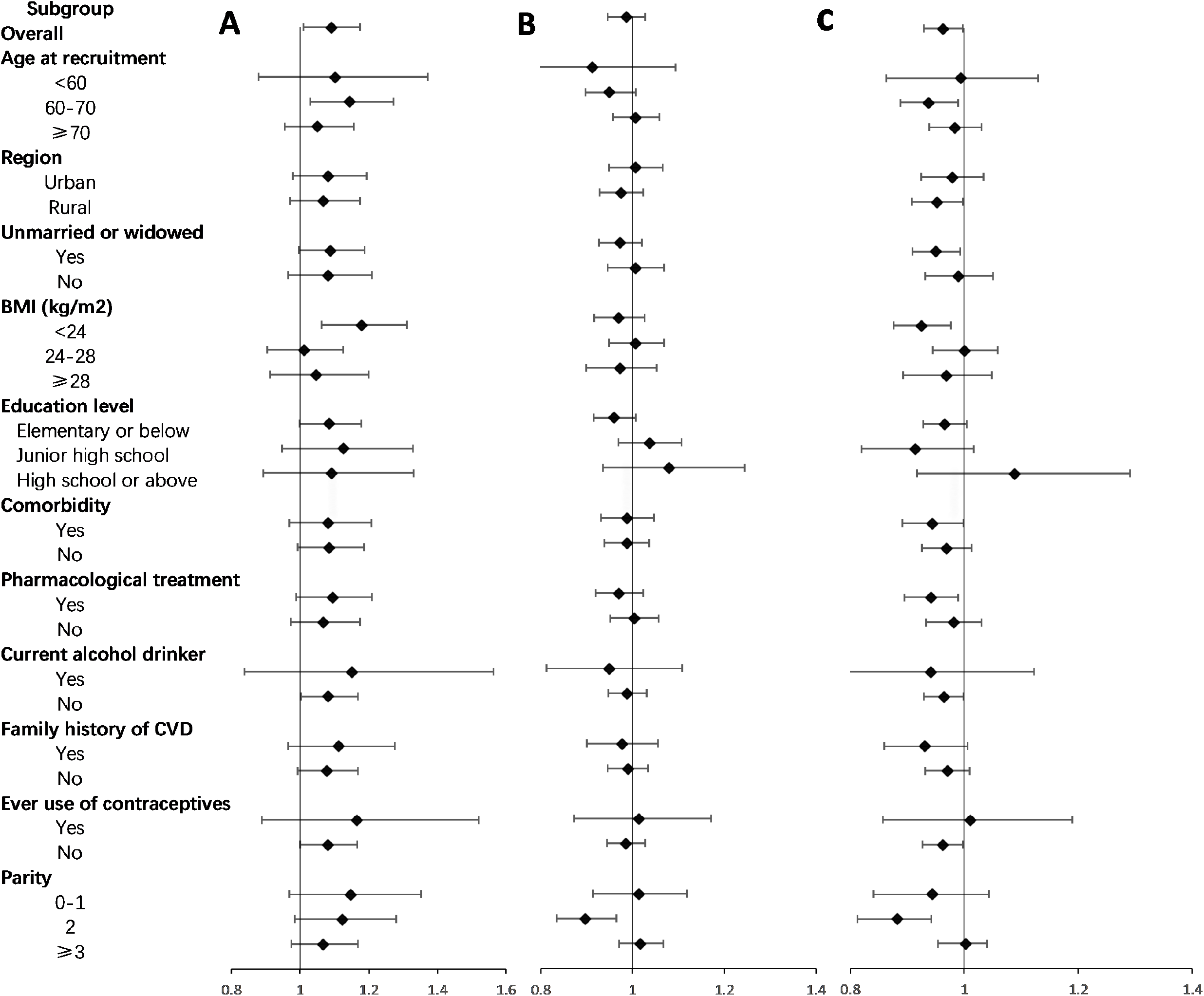
Adjusted hazard ratios (HRs) for stroke per year by age at menarche, age at menopause, and reproductive lifespan within various subgroups. A, adjusted HRs for age at menarche. B, adjusted HRs for age at menopause. C, adjusted HRs for reproductive lifespan. Model A were adjusted for age at recruitment, region, marital status, body mass index, education level, alcohol drinking, smoking, comorbidity, pharmacological treatment, family history of stroke, menstrual status, contraceptive use status, and breastfeeding experience, parity. Model B were adjusted for age atrecruitment, region, marital status, body mass index, education level, alcohol drinking, smoking, comorbidity, pharmacological treatment, family history of stroke, age at menarche, reproductive lifespan, contraceptive use status, and breastfeeding experience, parity. Model C were adjusted for age at recruitment, region, marital status, body mass index, education level, alcohol drinking, smoking, comorbidity, pharmacological treatment, family history of stroke, age at menarche, age at menopause, contraceptive use status, and breastfeeding experience, parity.

## 4. Discussion

In this study, we firstly evaluated multiple subgroups using a nationally prospective sample to comprehensively investigate the associations between age at menarche and menopause, reproductive lifespan, and risk of stroke events in Chinese women. We found that older menarche and shorter reproductive years were related to an increased risk of stroke in Chinese women. The association was independent of age, BMI, alcohol consumption and contraceptive pill use. Improvement in stroke awareness in women of childbearing age is required. Several studies have previously explored the association between age of menarche and stroke, but reported inconsistent results. The UK Million Women Study showed a significant U-shaped relationship between age at menarche and the outcomes stroke [14]. One cohort study reported that women with menarche age ≤13 years had a higher risk of ischemic stroke than those with ≤15 years, in which the participants were selected from a rural town in Japan[15]. In contrast, a study of 267,400 female textile workers in Shanghai found that early menarche was not associated with an increased risk of stroke mortality [16]. A case-control study reported that a higher age of menarche was associated with an increased risk of ischemic stroke, which was consistent with our study [17]. Part of the reason for this difference may be a small sample sizes,geographical limitations of a city or province, or only rural areas, age or ethnic differences, or residual confounding of unmeasured or other potential risk factors.

In addition, the relationship between age at menopause and stroke risk has been contradictory in previous studies. Early menopause was positively associated with coronary heart disease and stroke in a longitudinal, multi-ethnic cohort study independent of traditional CVD risk factors [18], which might have certain limitations about extrapolation due to survival bias. Two meta-analysis indicated women with early menopausal age had a higher risk of stroke morbidity and mortality [19, 20]. However, several studies found no convincing relationship between early menopausal age and CVD[16, 21-25], which were consistent with our findings. Differences in population and study design, as well as the influence of confounding factors, may be the leading reasons for the inconsistent results.

Previous studies investigating the association between reproductive lifespan and stroke risk were inconsistent. Results from the US National Health and Nutrition Examination Survey showed that in women aged 60 and older, longer reproductive lifespan were associated with a lower risk of stroke [26]. A multicenter case-controlstudy in Taiwan showed that longer reproductive lifespan was associated with a lower incidence of stroke in postmenopausal women [17]. A meta-analysis found that women with a shorter reproductive lifespan was associated with a 31% increased risk of stroke [27]. One cross-sectional study from the USA showed that an annual increase inreproductive lifespan was associated with 3% reduction in the risk of overall CVD and stroke events [26]. For stroke mortality, a large prospective study from Japan showed a reduced risk for longer reproductive lifespan [28]. An analysis of the Nurses’ Health Study found that a reproductive lifespan of 30 years or less was independently associated with an increased risk of total stroke [29]. These associations between reproductive lifespan and the risk of stroke are consistent with our study. However, some studies have found no significant association [24, 32-35] or even a U-shaped association [36]. Meanwhile, some studies have shown that there was a positive correlation betweenreproductive lifespan and the risk of stroke [30]. However, due to the lack of reproductive information, a representative prospective study is needed to verify the real association in the future.

Some mechanisms may explain the association between reproductive factors and risk of stroke. Studies have shown that late menarche is associated with low estrogen levels [31, 32]. Based on various experimental methods, estrogen has been shown to affect hemorrhage volume, tissue survival, cerebral blood flow, and the immune response, all of which may affect the response to stroke and improve outcomes [33]. Timing of menarche and menopause were related, and women who experienced early menarche were at higher risk of early menopause [29]. A longer reproductive lifespan with reduced risk of stroke can be attributed to prolonged exposure to endogenous estrogen [29]. In addition, changes in endogenous estrogens may affect lipid levels, contributing to alteration in the lipoprotein profiles that may subsequently lead to atherosclerosis [34]. One potential mechanism to explain these changes in lipoprotein profile is that large high-density lipoprotein particles are converted to smaller particles by increased hepatic lipase, and estrogen inhibits hepatic lipase [35]. Thus, thehypothesized mechanisms for these risks may be associated with shortened exposure time to beneficial endogenous estradiol and subsequent increases in higher-risk lipid profiles leading to atherosclerosis. These findings suggest that reproductive factors may play an important role in maintaining and improving cardiovascular health and could be used to assess populations with poor cardiovascular health for targeted interventions.

### 5. Strengths and limitations

Our study has important advantages: prospective design with standardized methods and strict quality control measures; adjustments for potential risk factors for stroke and detailed subgroup analysis to improve the reliability of results. There are several limitations to consider. First, age at menarche and menopause is known to vary by ethnicity [36-38]. However, it is unclear whether ethnicity plays a role in thisassociations. Since the study participants were all Chinese women, which minimized the confounding effects of ethnic background, but might reduce generalizations to otherraces. Secondly, ages at menarche and menopause were self-reported, which increases the possibility of misclassification due to recall bias. However, previous studies have shown that the recall of menarche [39, 40] and menopause age is relatively accurate [41, 42]. Self-reported were a reasonably effective and repeatable method, which would be non-differential and tend to weaken the true strengths of the associations. Lastly, although our paper has comprehensively adjusted for potential confounding factors, the possibility of influence of other residual factors cannot be completely ruled out.

## 6. Conclusion

Based on a national cohort study in Chinese women, we found that late age at menarche and shorter reproductive span were both significantly associated with increased risk of stroke. These associations also appeared to be similar amongst subgroups. In addition, there was a negative association between age at menopause and stroke risk in women with 2 live births. Our findings have important public health implications for early detection and timely implementation of appropriate interventions in women at high risk of stroke.

## Data Availability

The datasets used and/or analyzed during the current study are available from the corresponding author Zengwu Wang on reasonable request.

## Abbreviations

FPG: Fasting plasma glucose
TC: total cholesterol
TG: triglycerides
LDL-C: low-density lipoprotein cholesterol
HDL-C: high-density lipoprotein cholesterol
BP: blood pressure
BMI: Body mass index
SD: standard deviation
HRs: Hazard ratios
CIs: confidence intervals

## Table Legends

**Table 1. Characteristics of study participants by age at menarche**.

**Table 2. Characteristics of study participants by age at menopause**.

**Table 3. Characteristics of study participants by reproductive lifespan**.

## Declarations

## Ethics approval and consent to participate

Yes.

## Consent for publication

Yes.

## Sources of funding

The work was supported by the CAMS Innovation Fund for Medical Sciences [grant number 2017-I2M-1–004]; the China National Science & Technology Pillar Program [grant number 2011BAI11B01]; the surveillance of cardiovascular and its risk factors among Chinese Residents; he National Key R&D Program of China during the Thirteen Five-Year Plan Period [grant number No. 2018YFC1315303].

## Author statement

Chen Lu: Methodology, Formal analysis, Software, Writing an original draft preparation partly; Hu Zhen: Conceptualization, Writing an original draft preparation partly; Chen Zuo, Zhang Linfeng, Wang Xin and Zheng Congyi: Data curation, Formal analysis, Software; Cao Xue, Cai Jiayin, Song Yuxin, Tian Yixin, Huang Yilin and Gu Runqing: Investigation; Wang Zengwu: Conceptualization, Funding acquisition, Writing-Reviewing and Editing.

## Declaration of Competing Interest

None.

## Acknowledgments

This study was accomplished through the fine work of the staff at the national level, we thank all of colleagues involved in the survey and we also gratefully acknowledge Suning Li for maintaining the data. The authors are grateful to OMRON Corporation, Kyoto, Japan, for supporting the Blood Pressure Monitor (HBP-1300) and body fat and weight measurement device (Vbody HBF-371); Henan Huanan Medical Science & Technology Co., Ltd, China, for Digital ECG device (GY-5000); Microlife, Taipei, Taiwan, for Automated ABI device (Watch BP Office device.

## Notes

### Competing Interest Statement

The authors have declared no competing interest.

### Author Declarations

The Ethics Committee of Fuwai Hospital (Beijing, China) approved the study.

